# Identifiability of parameters in mathematical models of SARS-CoV-2 infections in humans

**DOI:** 10.1101/2022.04.26.22274345

**Authors:** Stanca M. Ciupe, Necibe Tuncer

**Affiliations:** Department of Mathematics, Virginia Polytechnic Institute and State University, 225 Stanger Street, Blacksburg, VA, 24060 USA; Department of Mathematics, Florida Atlantic University, 777 Glades Road, Boca Raton, FL, 33431 USA

## Abstract

Determining accurate estimates for the characteristics of the severe acute respiratory syndrome coronavirus 2 in the upper and lower respiratory tracts, by fitting mathematical models to data, is made difficult by the lack of measurements early in the infection. To determine the sensitivity of viral predictions to the noise in the data, we developed a novel two-patch within-host mathematical model and investigated its ability to match population level data. We proposed several approaches that can improve practical identifiability of parameters, including an optimal experimental approach, and found that availability of viral data early in the infection is of essence for improving the accuracy of the estimates. Our findings can be useful for designing interventions.

## Introduction

Understanding the upper respiratory tract (URT) kinetics of the severe acute respiratory syndrome coronavirus 2 (SARS-CoV-2) is important for designing public health interventions such as testing, isolation, quarantine, and drug therapies [12,13,16,17,22–25,30,35,41,42]. Similarly, understanding the kinetics of SARS-CoV-2 in the lower respiratory tract (LRT) is important for predicting the potential for severe disease, respiratory failure, and/or death [8, 24]. Insights into the mechanism of SARS-CoV-2-host interactions and their role in transmission and disease have been found using mathematical models applied to longitudinal data [16, 17, 21–25, 30, 35, 41, 42]. While these studies are instrumental in determining important parameters (such as SARS-CoV-2 daily shedding and clearance rates, basic reproduction number, the role of innate immune responses in controlling and/or exacerbating the disease), their predictions are limited by the lack of data early in the infection. As such, with few (if any) samples available before viral titers peak, the early virus kinetics and the mechanisms for these early kinetics are uncertain. In this study, we investigate the sensitivity of the predicted outcomes of a within-host model of SARS-CoV-2 infection to the availability of data during different stages of the infection and use our findings to make recommendation.

A German study by Wolfel *et al*. collected data from nine patients infected early in the pandemic through contact with the same index case [44]. The study showed independent virus replication in upper and lower respiratory tracts [7,44] suggesting the possibility that virus kinetics, disease stages, and host involvement in control and pathogenesis are dependent on which area of the respiratory tract is homing SARS-CoV-2 at different stages of the disease [27, 33, 37]. One shortcoming when evaluating the data in this study comes from the fact that viral RNA was collected only after the patients became symptomatic, with an estimated first data point available on average 5-7 days after infection. Several within-host mathematical models developed and applied to the data set in the Wolfel *et al*. study have evaluated SARS-CoV-2 parameters, determined the role of innate immune responses, found connections between total RNA and infectious titers, and identified the efficacy of drug therapies [22, 24, 42]. We are interested in determining how the lack of data early in the infection affects these estimates.

We first developed our own within-host model that does not consider innate immunity explicitly and used the data from Wolfel *et al*. to estimate pertinent parameters. We next investigated the sensitivity of the estimated parameters to the presence of data at different stages of infection. To accomplish this, we created virtual data sets that span various stages of the infection and determined how our initial predictions are being influenced by the additional data. Such results may influence our understanding of both viral expansion and the effect of inoculum dose on disease progression.

## Methods

### Mathematical Model

SARS-CoV-2 virus infects and replicates in epithelial cells of the upper and lower respiratory tract [44]. We model this by developing a two patch within-host model, where the patches are the two respiratory tracts which are linked through viral shedding. Both respiratory tract patches assume interactions between uninfected epithelial cells, *T*_*j*_; infected epithelial cells, *I*_*j*_; and virus homing in tract *j, V*_*j*_ at time *t*. Here, *j* = {*u, l*}, with *u* describing the URT patch and *l* describing the LRT patch. Target cells in each patch get infected at rates *β*_*j*_ and infected cells produce new virions at rates *p*_*j*_. Infected cells die at rates *δ*_*j*_ and virus particles are cleared at a linear rate *c*_*u*_ in the upper respiratory tract and in a density dependent manner *c*_*l*_*V*_*l*_*/*(*V*_*l*_ + *K*) in the lower respiratory tract. The two patches are linked via the virus populations, with a proportion *k*_*u*_ of *V*_*u*_ migrating from URT to LRT and *k*_*l*_ of *V*_*l*_ migrating from LRT to URT. The model describing these interactions is given by

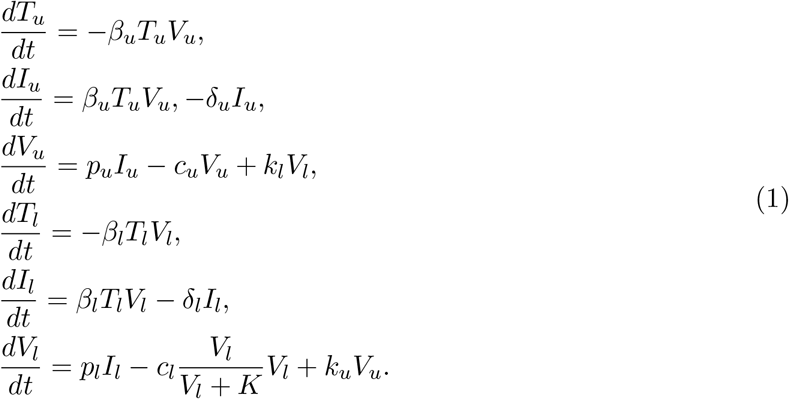

We model the initial conditions of the model Eq. 1 as follows. We assume that all epithelial cells in the URT and LRT patches are susceptible to virus infection. When infection occurs, it results in a small initial virus inoculum which homes in the URT alone. Under these assumptions, system Eq. (1) is subject to initial conditions

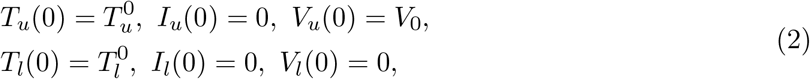

where *V*_0_ is the viral inoculum. We aim to determine the dynamics of system Eq. (1) over time for model parameters that explain URT and LRT tract data in a single patient (patient A) and in the population data (all nine patients) from [44].

### Parameter Estimation

#### Patient Data

In January 2020, nine patients tested positive for COVID-19 in a single-source outbreak in Bavaria, Germany [7]. Early detection allowed for rapid contact tracing, testing, and monitoring of the affected community: young healthy professionals in their mid-thirties. A followup study published time series for the post symptoms virus data isolated from oral-and nasopharyngeal throat swabs (in copies per swabs) and from sputum samples (in RNA copies per mL) for the same patient population over their entire course of disease. The patients’ throat swabs and sputum data (Figure 2 of [44]) were obtained through personal communication with the authors. Since we know the incubation period for each patient [7] (see Table 1), we assume time zero in our study to be the day of infection for the patients in [44].

**Table 1:**
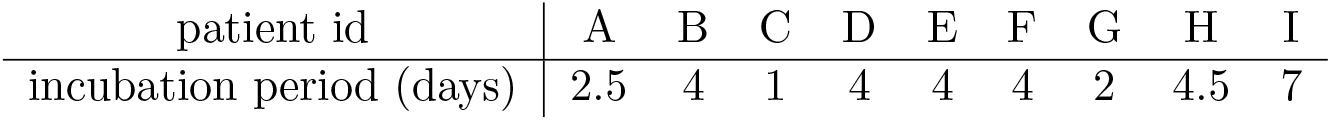
Incubation periods estimated in [7].

#### Identifiability Analysis

Using the URT and LRT viral load data, we aim to determine the unknown parameters ***p*** = {*β*_*u*_, *δ*_*u*_, *p*_*u*_, *c*_*u*_, *k*_*l*_, *β*_*l*_, *δ*_*l*_, *p*_*l*_, *c*_*l*_, *K, k*_*u*_} of the within-host model Eq. (1). Before attempting to estimate the within-host model parameters using noisy laboratory data, it is crucial to analyze whether the model is structurally identifiable. Specifically, we need to know if the within-host model Eq. (1) is structured to reveal its parameters from upper and lower viral load observations. We approach this problem in an ideal setting where we assume that the observations are known for every *t >* 0 and they are not contaminated with any noise. This analysis is called structural identifiability [18].

The observed data in Wolfel *et al*. [44] is modeled in the within-host model Eq. (1) by variables *V*_*u*_ and *V*_*l*_, which account for the upper and lower respiratory tract viral titers. We denote these observed variable as

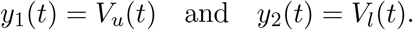

First, we give the definition of structural identifiability in terms of the observed variables *y*_1_(*t*) and *y*_2_(*t*) [10, 18, 38, 39].

##### Definition 1

*Let* ***p*** *and* ***q*** *be the two distinct vectors of within-host model Eq. (1) parameters. We say that the within-host model is structurally (globally) identifiable if and only if*

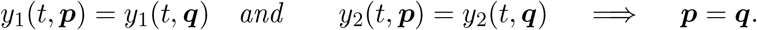

Simply put, we say that the within-host model Eq. (1) is structurally identifiable if two identical observation are only possible for identical parameters. We perform the structural identifiability analysis via differential algebra approach. The first step in this approach is eliminating the unobserved state variables from the within-host model Eq. (1). The reason for eliminating the unobserved state variables is to obtain a system which only involves the observed states and model parameters. Since this is a complex procedure, we use DAISY [5] and obtain the following system

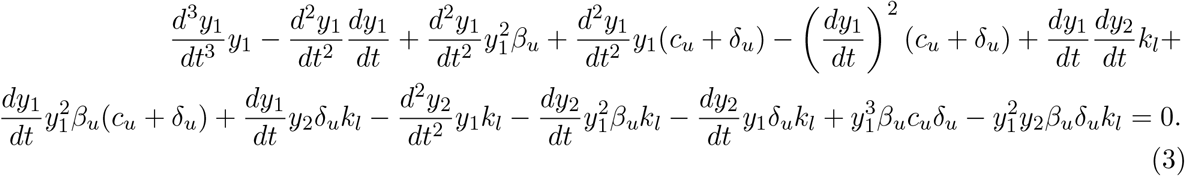

and

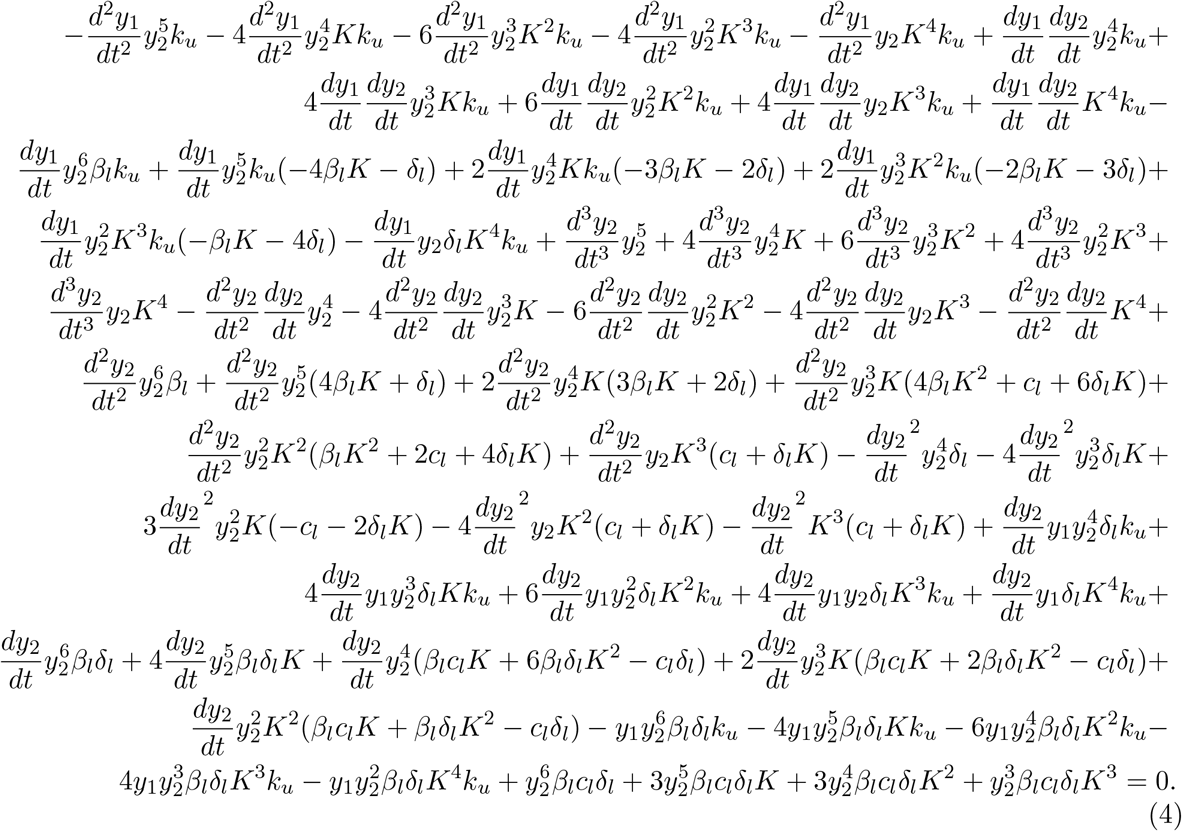

Eq. (3) and Eq. (4) are called input-output equations of within-host model Eq. (1), which are differential polynomials involving the observed state variables *y*_1_ = *V*_*u*_(*t*) and *y*_2_ = *V*_*l*_(*t*) and the within-host model parameters. Note that solving input-output equations Eq. (3) and Eq. (4) is equivalent to solving the within-host model Eq. (1) for the state variables *V*_*u*_(*t*) and *V*_*l*_(*t*). For identifiability analysis, it is crucial that the input-output equations are monic, *i*.*e*. the leading coefficient is 1. It is clear that the input-output equation Eq. (3) is monic, and the input-output equation Eq. (4) can be made monic by dividing the coefficients with the coefficient of the leading term, which is *k*_*u*_. As a result, the definition of the structural identifiability within differential algebra approach which involves input-output equations takes the following form [10, 18, 38, 39].

##### Definition 2

*Let c*(***p***) *denote the coefficients of the input-output equations*, (3) *and* (4) *where* ***p*** *is the vector of model parameters. We say that the within-host model Eq. (1) is structured to reveal its parameters from the observations if and only if*

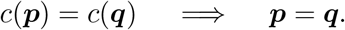

Suppose ***p*** = {*β*_*u*_, *δ*_*u*_, *p*_*u*_, *c*_*u*_, *k*_*l*_, *β*_*l*_, *δ*_*l*_, *p*_*l*_, *c*_*l*_, *K, k*_*u*_} and 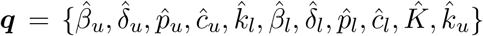 are two parameter sets of the within-host model which produced the same observations. This can only happen if the coefficients of the input-output equations Eq. (3) and Eq. (4) are the same. Hence, if *c*(***p***) denote the coefficients of the corresponding monic polynomial of input-output equations, we solve *c*(***p***) = *c*(***q***) to obtain

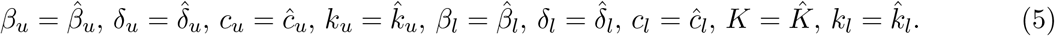

The solution set (5) means that the parameters, *β*_*u*_, *δ*_*u*_, *c*_*u*_, *k*_*u*_, *β*_*l*_, *δ*_*l*_, *c*_*l*_, *K* and *k*_*l*_ can be identified uniquely. However, parameters *p*_*u*_ and *p*_*l*_ both disappear from the input-output equations Eq. (3) and Eq. (4). It is easier to see the reason behind this by scaling the unobserved state variables of the within-host model Eq. (1) with a postive scalar *σ* > 0. Hence, 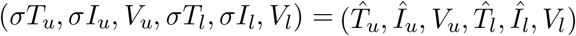 will solve the following system

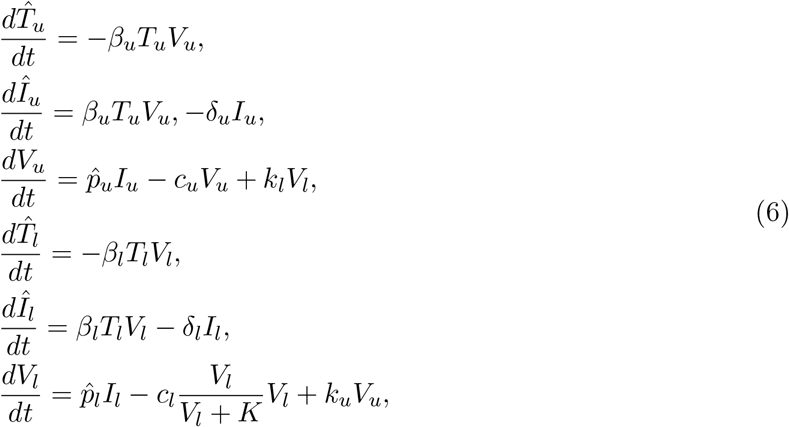

where 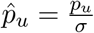 and 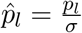. Since *σ* > 0 was arbitrary and the observations do not give information about the scaling parameter *σ*, the parameters *p*_*u*_ and *p*_*l*_ can not be identified from the viral load in the URT and LRT tracts. We conclude that the within-host model Eq. (1) is not identifiable. We summarize the structural identifiability result in the following Proposition 1.

##### Proposition 1

*The within-host model Eq. (1) is not structured to reveal its parameters from the observations of viral load in upper and lower respiratory tracts. The parameters p*_*u*_ *and p*_*l*_ *are not identifiable and only the parameters β*_*u*_, *δ*_*u*_, *c*_*u*_, *k*_*u*_, *β*_*l*_, *δ*_*l*_, *c*_*l*_, *K, k*_*l*_ *can be identified*.

To obtain a structurally identifiable model from the *V*_*u*_ and *V*_*l*_ observations, we scale the unobserved state variables with 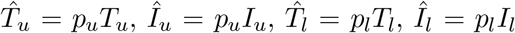 and obtain the following scaled within-host model

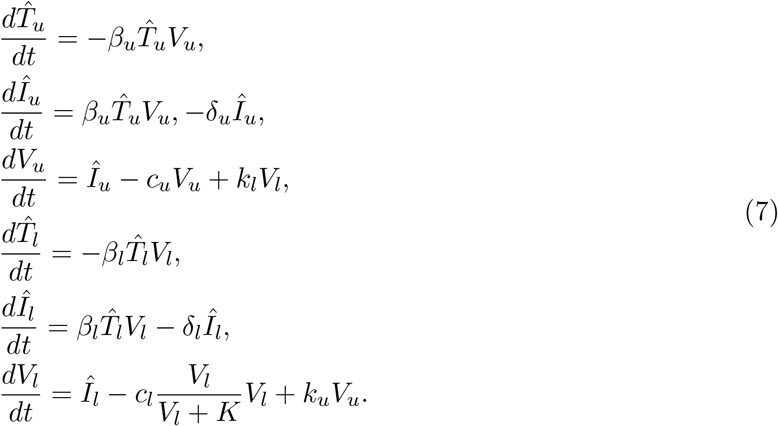

##### Proposition 2

*The scaled within-host model Eq. (7) is structured to reveal its parameters from the observations of viral load in upper and lower respiratory tracts. All the parameters*

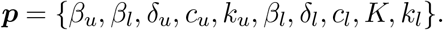

*can be identified, hence the within-host model Eq. (7) is globally identifiable*.

## Data fitting

### Parameter values

We assume that the upper respiratory tract susceptible population is 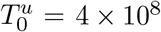 epithelial cells, as in influenza studies [2]. This estimate was obtained by assuming a URT surface in adults of 160cm^2^ [28] and an epithelial cell’s surface area of 2 × 10^*−*11^ − 4 × 10^*−*11^m^2^ [11]. We use a similar method to estimate the target cell population in the LRT. The lung”s surface area is 70m^2^ (with range between 35m^2^ and 180m^2^) [15] is composed of gas exchange regions (aveoli), and of conducting airways (trachea, bronchi, bronchioles). Since the gas exchange region is affected by SARS-Cov-2 only in severe cases [27] we ignore it, and restrict the LRT compartment to the conducting airways whose surface area is 2471 ± 320cm^2^ [29]. Therefore, we obtain an initial epithelial cell target population in the LRT of 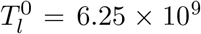 pithelial cells. If we assume that viral production rates are *p*_*u*_ = 50 and *p*_*l*_ = 32 per day then, after scaling, we have initial target cell populations in the URT and LRT of 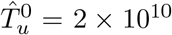 epithelial cells and 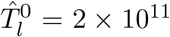 epithelial cells. The other initial conditions are unaffected by scaling and 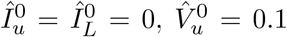 and 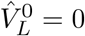, where the virus inoculum of 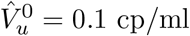 is set below the reported limit of quantification of 10^2^ cp/ml [44]. Lastly, the incubation periods were estimated in [7] and are listed in Table 1.

### Bayesian parameter estimation

During the data collection process, observations are perturbed with noise. Hence, the URT and LRT viral load deviates from the smooth trajectory of the observations *y*_1_(*t*) and *y*_2_(*t*) at measurement times. We represent measurement error using the following statistical model

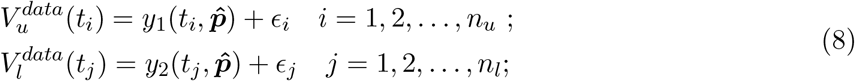

where 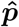 is the true parameter vector assumed to generate the data, and the random variables *ϵ*_*I*_ and *ϵ*_*j*_ are assumed to be Gaussian with mean zero and standard deviation *σ*.

We use Bayesian inference and Markov Chain Monte Carlo (MCMC) to determine the remaining nine parameters of the model Eq. (7)

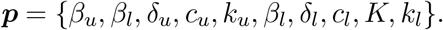

Bayesian inference treats model parameters as random variables and seeks to determine the parameters” posterior distribution, where the term “posterior” refers to data-informed distributions. The posterior densities are determined using Bayes” theorem, which defines them as the normalised product of the prior density and the likelihood. Let *π*(***p***|𝒟) denote the probability distribution of the parameter ***p*** given the data 𝒟 = (*V*_*u*_(*t*_*i*_), *V*_*l*_(*t*_*j*_)), then the Bayes theorem states that

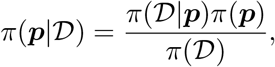

where *π*(***p***) is the prior parameter distribution and *π*(𝒟) is a constant which is usually considered to be a normalization constant so that the posterior distribution is indeed a probability density function (pdf), *i*.*e*. its integral equals to 1. The likelihood function *π*(𝒟|***p***) gives the probability of observing the measurements D given that the parameter values is ***p***. In terms of the within-host model Eq. (7) and the laboratory data Eq. (8), the likelihood function *π*(𝒟|***p***) takes the following form

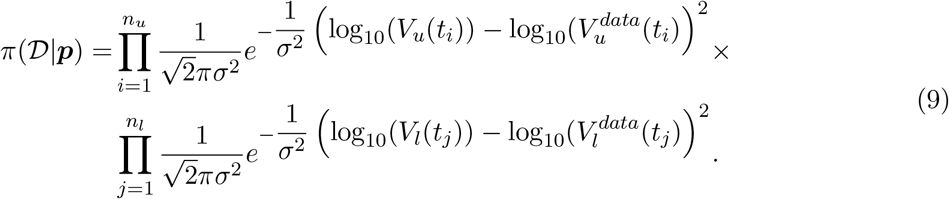

The ultimate goal is to determine the posterior distributions of the parameters in the light of laboratory data. To approximate the posterior distributions, we use the MCMC method introduced in [19, 20]. MCMC methods generate a sequence of random samples ***p***_1_, ***p***_2_, …, ***p***_*N*_ whose distribution asymptotically approaches the posterior distribution for size *N*. The random walk Metropolis algorithm is one of the most extensively used MCMC algorithms. The Metropolis algorithm starts at position ***p***_*i*_, then the Markov chain generates a candidate parameter value ***p****∗* from the proposal distribution, and the algorithm accepts or rejects the proposed value based on probability *α* given by

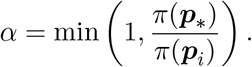

As with the Metropolis algorithm, the essential feature of MCMC approaches is the formulation of a proposal distribution and an accept-reject criteria. In this paper, we employ the Delayed Rejection Adaptive Metropolis, (DRAM [19]) and use the MATLAB toolbox provided by the same authors [26]. In comparison to other Metropolis algorithms, the Markov chain constructed with DRAM is robust and converges rapidly (see Figure 1).

**Figure 1:**
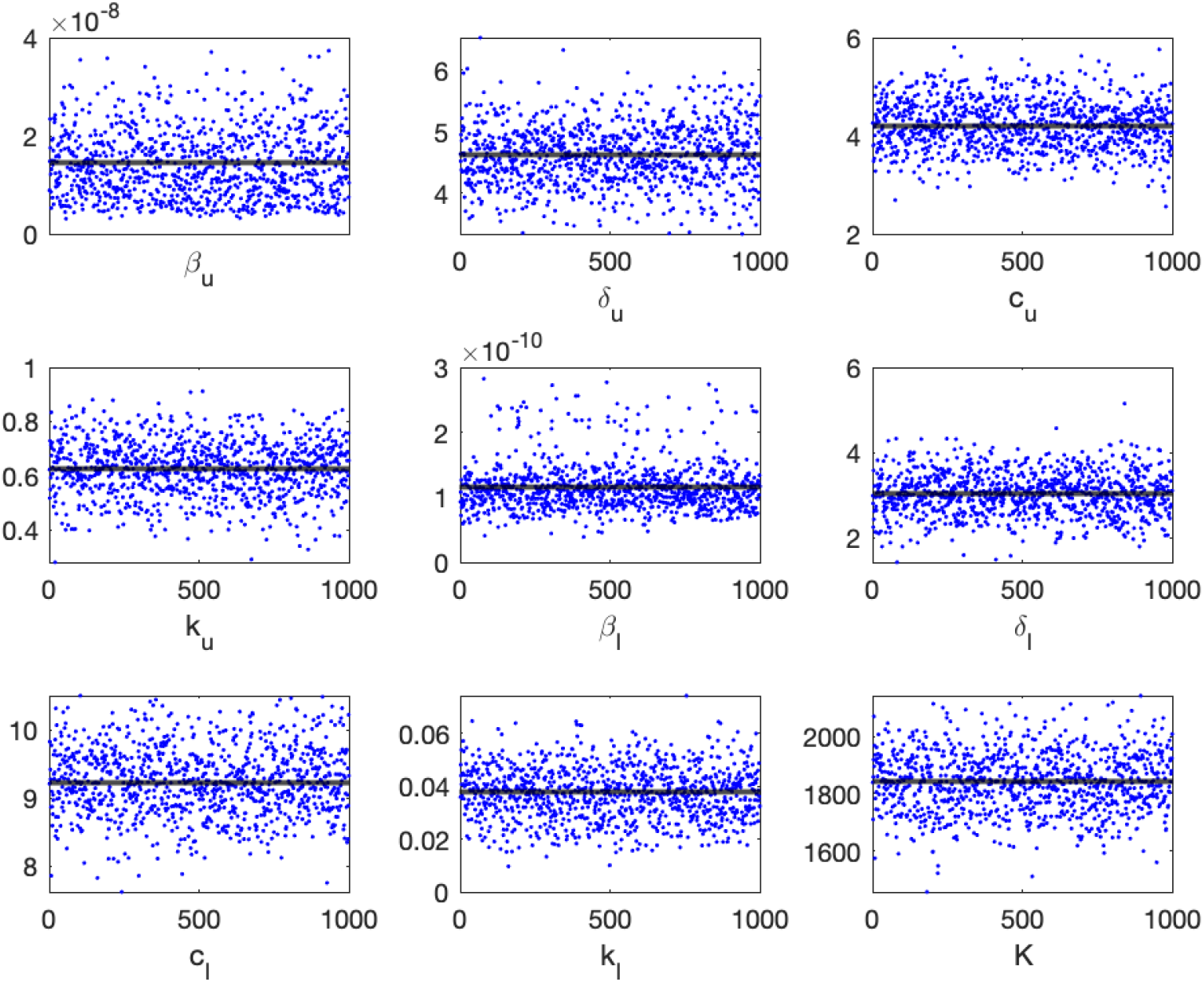
The Markov chain of the within-host model Eq. (7)’s parameters obtained when the model is fitted to the population data. Every 1000^*th*^ point of 10^6^ iterations are shown. The black line shows the mean of the chain.

The two patch within-host model Eq. (7) is novel, hence we do not have any prior information regarding model parameters. We determine the prior distributions by fitting the structurally identifiable within-host model Eq. (7) to patient A’s data and to the population data (all nine patients). The prior distributions *π*(***p***) are then defined as a normal distribution with a mean equal to the fitted value and variance equal to *σ*^2^, *π*(***p***) ∼ *N* (***µ, σ***). Table 2 shows the obtained prior distribution of each parameter for patient A and population data, together with the lower and upper bounds of the prior *π*(***p***).

**Table 2:** Parameters for the within-host model Eq. (7) are listed together with their lower and upper bounds for the priors. Prior distributions are normally distributed with mean equal to the fitted value and variance, *σ*^2^.

| Parameter | Description | Prior $\pi(\mathbf{p})$<br>Min-Max | Patient A<br>$\pi(\mathbf{p}) \sim \mathcal{N}(\boldsymbol{\mu}, \boldsymbol{\sigma})$ | Population<br>$\pi(\mathbf{p}) \sim \mathcal{N}(\boldsymbol{\mu}, \boldsymbol{\sigma})$ |
| --- | --- | --- | --- | --- |
| $\beta_u$ | viral infectivity in URT | $(10^{-12}, 10^{-7})$ | $\mathcal{N}(1.1 \times 10^{-8}, 10^{-8})$ | $\mathcal{N}(8.9 \times 10^{-9}, 10^{-8})$ |
| $\beta_l$ | viral infectivity in LRT | $(10^{-12}, 10^{-7})$ | $\mathcal{N}(3.9 \times 10^{-8}, 10^{-10})$ | $\mathcal{N}(9.3 \times 10^{-11}, 10^{-10})$ |
| $\delta_u$ | infected cell decay rate in URT | $(0, 50)$ | $\mathcal{N}(4.88, 0.5)$ | $\mathcal{N}(4.64, 0.5)$ |
| $\delta_l$ | infected cell decay rate in LRT | $(0, 50)$ | $\mathcal{N}(5.59, 0.5)$ | $\mathcal{N}(2.99, 0.5)$ |
| $c_u$ | viral decay rate in URT | $(0, 30)$ | $\mathcal{N}(2.88, 0.5)$ | $\mathcal{N}(4.27, 0.5)$ |
| $c_l$ | viral decay rate in LRT | $(0, 30)$ | $\mathcal{N}(11.43, 0.5)$ | $\mathcal{N}(9.21, 0.5)$ |
| $K$ | $V_u$ half-maximal viral loss | $(0, 3000)$ | $\mathcal{N}(910, 100)$ | $\mathcal{N}(1840, 100)$ |
| $k_u$ | shedding into LRT | $(10^{-6}, 1)$ | $\mathcal{N}(0.24, 0.1)$ | $\mathcal{N}(0.62, 0.1)$ |
| $k_l$ | shedding into URT | $(10^{-6}, 1)$ | $\mathcal{N}(0.0008, 0.0001)$ | $\mathcal{N}(0.036, 0.01)$ |

## Results

### Viral dynamics

To study the kinetics of SARS-CoV-2 in the upper and lower respiratory tracts we developed a two patch within-host model Eq. (1) that assumed viral shedding between the two patches. To ensure structural identifiability, we rescaled our equations by removing the non-identifiable parameters *p*_*u*_ and *p*_*l*_ (see *Identifiability Analysis* section in *Material and Methods*). The resulting model Eq. (7) was validated against SARS-CoV-2 RNA data from throat swabs and sputum samples collected from an infectious event with the same index case early in the pandemic [44]. We used Bayesian parameter estimation with the viral samples in URT and LRT from a single individual (patient A) and the entire population (nine individuals) and approximated posterior distributions with *N* = 10^6^ Markov chain iterations (see *Data fitting* section in *Material and Methods*).

We generated prediction graphs of the within-host model Eq. (7) by sampling parameter realizations from posterior distributions. The model’s predictive posterior distribution for single patient URT-LRT viral data and population URT-LRT viral data are presented in Figure 2. The resulting dynamics show viral expansion to peak values at days 2.1 in URT and 2.9 in LRT followed by decline in both tracts (see Figure 2). The grey areas in the graph represent the 50% and 95% posterior regions. The fewer data points in patient A results in wider model prediction range (gray regions) compared to the population predictions, especially for the LRT viral load.

**Figure 2:**
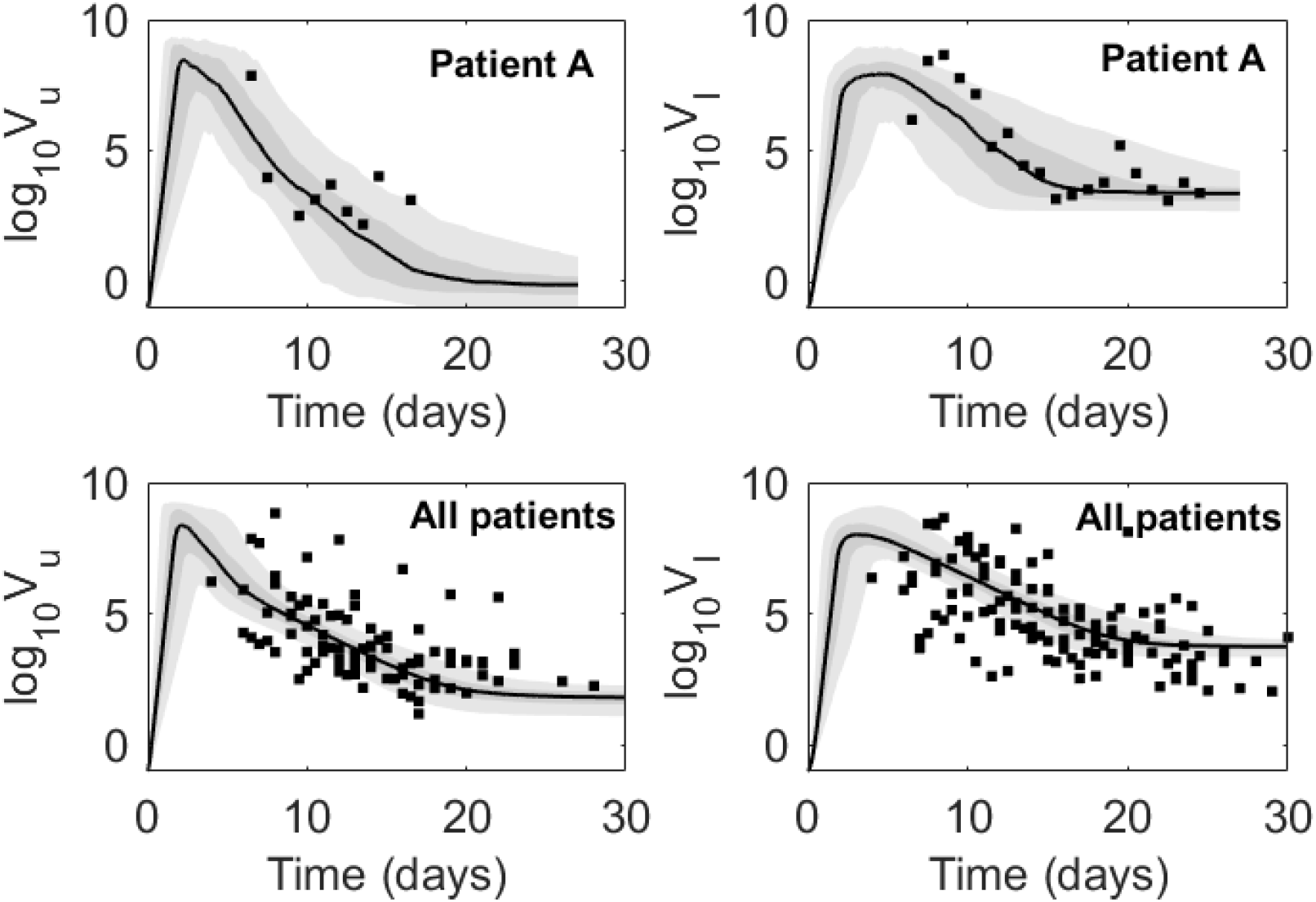
Virus dynamics obtained from fitting within-host model Eq. (7) to URT virus titer (left) and LRT virus titer (right) in patient A and in the entire population. The grey bars represent 50% and 95% posterior regions.

While the viral titers decay to low levels (below 10^2^ cp/ml) three weeks after infection in the URT, they stay elevated (to above 5.4 × 10^3^ cp/ml at week four) in the LRT. To model viral RNA persistence in the LRT we included a density dependent term for the loss of LRT virus, *c*_*l*_*V*_*l*_*/*(*V*_*l*_ + *K*), and estimated parameter *K* where *V*_*l*_ loss is half-maximal, together with the other viral specific terms.

We found similar mean infectivity rates in the URT for both the individual patient considered (patient A) and the entire population, *β*_*u*_ = 1.4 × 10^*−*8^ ml/(vir× day). By contrast, the mean infectivity rates in the LRT for patient A is 3.2-times higher than the LRT infectivity rate of the total population, *β*_*l*_ = 3.9 × 10^*−*10^ ml/(vir× day) versus *β*_*l*_ = 1.2 × 10^*−*10^ ml/(vir× day). The mean infected cells death rates are similar in URT and LRT, *δ*_*u*_ = 4.9, *δ*_*u*_ = 4.6 per day and *δ*_*l*_ = 5.7, *δ*_*l*_ = 3 per day for patient A and for the total population, respectively. The mean viral clearance rates are higher in LRT compared to URT, *c*_*l*_ = 11.5, *c*_*l*_ = 9.2 per day compared to *c*_*u*_ = 2.8, *c*_*u*_ = 4.2 per day, for patient A and for the total population, respectively. This may indicate increased immune responses occurring in LRT. The mean URT to LRT shedding rates are higher than the mean LRT to URT shedding rates, *k*_*u*_ = 0.24, *k*_*u*_ = 0.63 (swab/ml) per day compared to *k*_*l*_ = 7.9 × 10^*−*4^, *k*_*l*_ = 0.04 (ml/swab) per day for patient A and for the total population, respectively. This one way shedding was observed by other studies that investigated the Wofle *et al*. data [24]. Lastly, the mean LRT viral load where viral clearance is half-maximal is *K* = 910 RNA per ml for patient A and *K* = 1841 RNA per ml for the total population.

### Practical identifiability

During MCMC data fitting, we used parameters limits predetermined to range around a single point estimation obtained using the “fminsearch” algorithm in Matlab (see Table 2). Parameter distributions for the nine parameter considered ***p*** = {*β*_*u*_, *β*_*l*_, *δ*_*u*_, *c*_*u*_, *k*_*u*_, *β*_*l*_, *δ*_*l*_, *c*_*l*_, *K, k*_*l*_} were obtained using an MCMC Bayesian approach that sampled the parameter space *N* = 10^6^ times. We apply DRAM MCMC algorithm and observe fast convergence of the chains (see Figure 1). The resulting distributions, together with the prior probability density functions (pdf) are presented in Figure 3. We observe good agreement between the prior pdf and the posterior distributions for all parameters with the exception of infectivity rates *β*_*u*_ and *β*_*l*_. Moreover, while all parameters follow normal distributions for patient A (Figure 3A), the LRT infectivity rate *β*_*l*_ follows a bimodal distribution in the fit to the total population data (Figure 3B).

**Figure 3:**
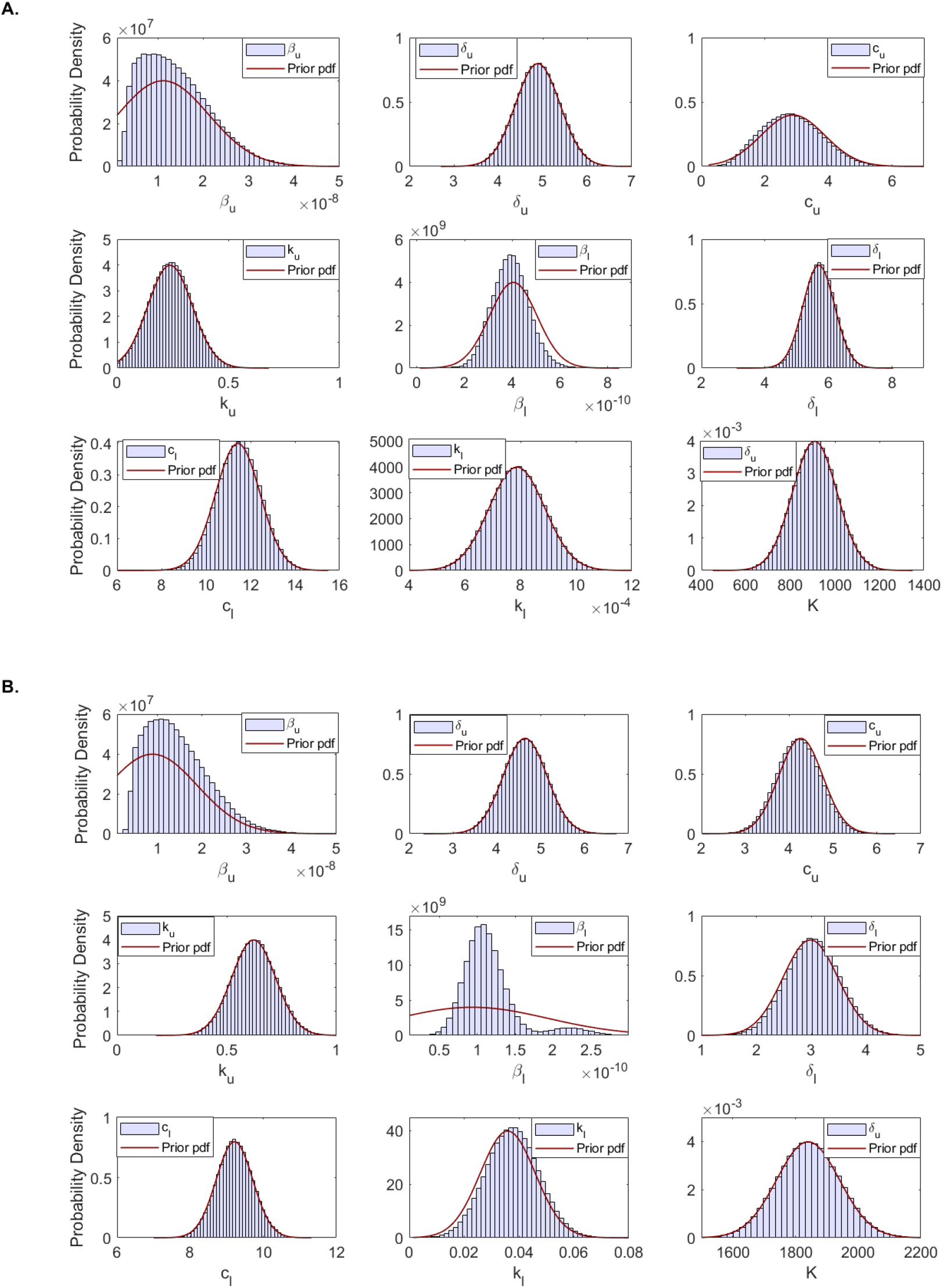
Histogram of estimated parameter distributions from fitting within-host model Eq. (7) to URT virus titer and LRT virus titer in: (A.) patient A and (B.) entire population. All parameters were considered normally distributed.

Figure 4 shows the scatter plots of for paired (*β*_*l*_, *k*_*l*_), (*β*_*l*_, *K*), (*β*_*l*_, *δ*_*l*_) and (*β*_*l*_, *c*_*l*_) parameter distributions obtained when the within-host model Eq. (7) is fitted to patient A’s data (panel A) and population data (panel B) (see also Figure S2 for the scatter plots of all parameter distributions). In the scatter plots for the population data containing parameter *β*_*l*_ we observe bimodal clustering. In joint density plots, bimodal clustering may suggest practical unidentifiability [36]. This suggests that, despite the fact that we have shown that the within-host model Eq. (7) is structurally identifiable, it may in fact not be practically unidentifiable. It is well understood that a structurally identifiable model may be practically unidentifiable [10,38–40]. Many variables can lead to practical unidentifiability, such as considerable noise in the data, a lack of enough data points, or timing of data collection.

**Figure 4:**
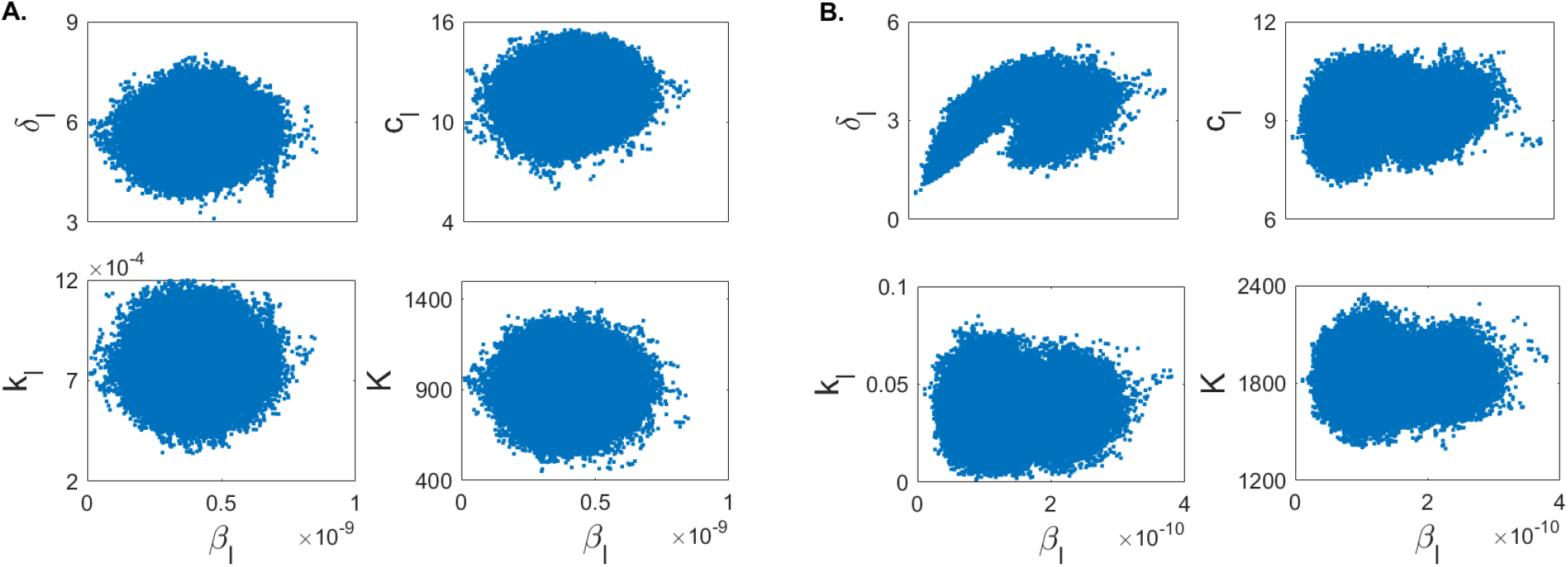
Scatter plots showing correlation among relevant parameters for (A.) patient A and (B.) total population.

### Optimal experimental design

The possible lack of practical identifiability for the total population may be due to (1) restrictions on the parameter space and the types of distributions we are imposing on the parameters, or (2) the limited data points early in the infection.

To investigate the first hypothesis, we collected samples in the parameter space of

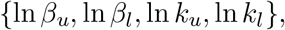

rather than {*β*_*u*_, *β*_*l*_, *k*_*u*_, *k*_*l*_} and the assumed that either {ln *β*_*u*_, ln *β*_*l*_, ln *k*_*u*_, ln *k*_*l*_} are normally distributed, or that {*β*_*u*_, *β*_*l*_, *k*_*u*_, *k*_*l*_} are lognormally distributed. We set the limits of logarithmic parameter priors as in Table 3 while keeping the limits of the other parameters as before (see Table 2). We sampled the new parameter space *N* = 10^6^ times and reapplied the MCMC Bayesian approach. The resulting estimates for parameters ***p*** = {*β*_*u*_, *β*_*l*_, *δ*_*u*_, *c*_*u*_, *k*_*u*_, *β*_*l*_, *δ*_*l*_, *c*_*l*_, *K, k*_*l*_} no longer show bimodal results regardless on whether we assume that {ln *β*_*u*_, ln *β*_*l*_, ln *k*_*u*_, ln *k*_*l*_} are normally distributed (see Figure 5A) or that {*β*_*u*_, *β*_*l*_, *k*_*u*_, *k*_*l*_} are lognormally distributed (see Figure 5B).

**Table 3:** Adjusted parameters for the within-host model Eq. (7) are listed together with their lower and upper bounds for the priors which are normally distributed with mean equal to the fitted value and variance, *σ*^2^.

| Parameter | Description | Prior $\pi(\mathbf{p})$<br>Min-Max | Population<br>$\pi(\mathbf{p}) \sim \mathcal{N}(\boldsymbol{\mu}, \boldsymbol{\sigma})$ |
| --- | --- | --- | --- |
| $\ln \beta_u$ | viral infectivity in URT (log scale) | $(-25, -14)$ | $\mathcal{N}(-18.5, 1)$ |
| $\ln \beta_l$ | viral infectivity in LRT (log scale) | $(-30, -15)$ | $\mathcal{N}(-23, 1)$ |
| $\ln k_u$ | shedding into LRT (log scale) | $(-11, 0)$ | $\mathcal{N}(-0.5, 1)$ |
| $\ln k_l$ | shedding into URT (log scale) | $(-7, 4)$ | $\mathcal{N}(-3.3, 1)$ |

**Figure 5:**
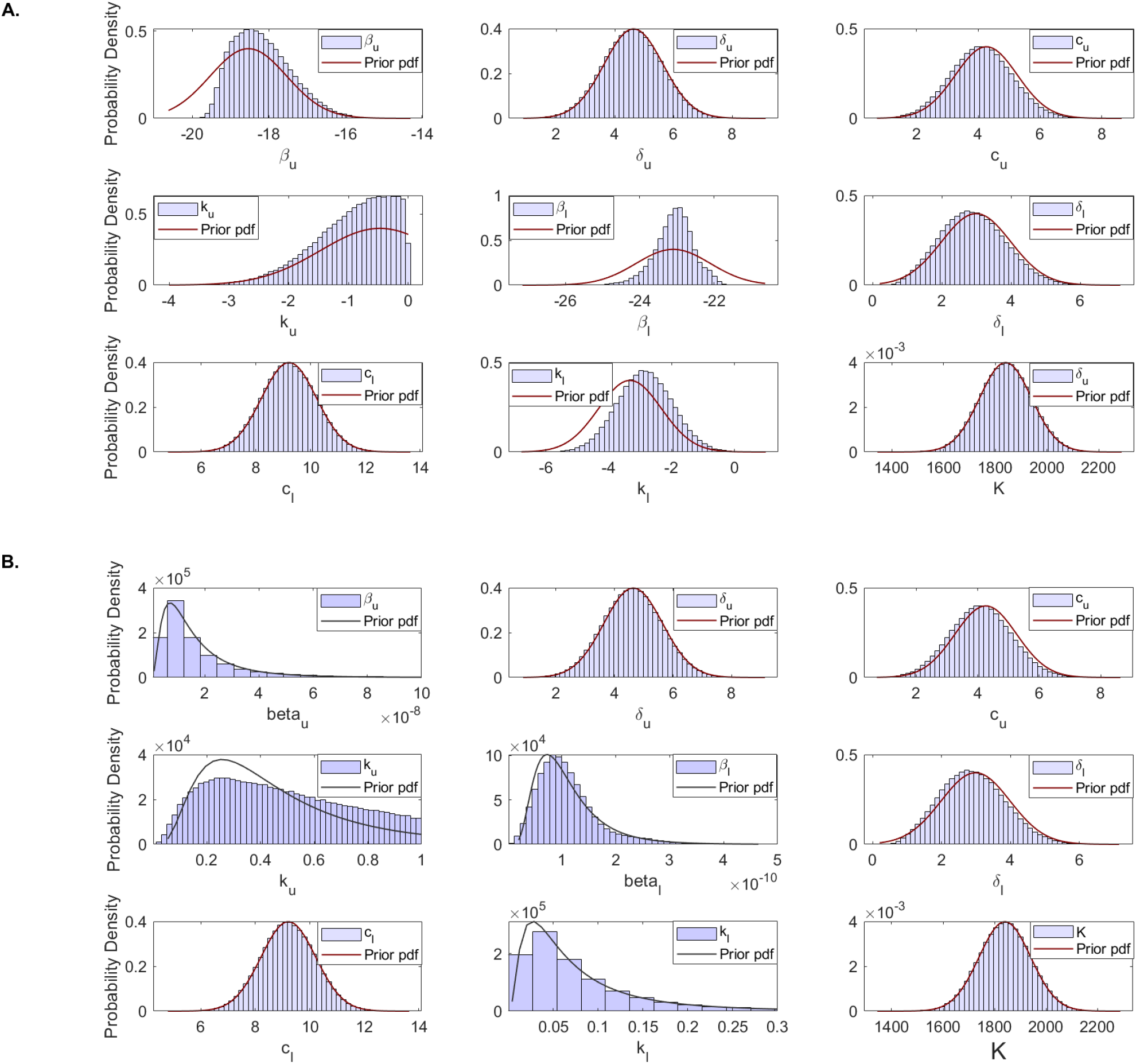
Histogram of estimated parameter distributions from fitting model Eq. (7) to URT virus titer and LRT virus titer in total populations. (A) Parameters ln *β*_*u*_, ln *β*_*l*_, ln *k*_*u*_, ln *k*_*l*_ were considered normally distributed. (B.) Parameters *β*_*u*_, *β*_*l*_, *k*_*u*_, *k*_*l*_ were considered lognormally distributed. All other parameters were considered normally distributed.

To investigate the second hypothesis, we created synthetic data and used it to further examine how the timing of the data collection in the population correlates to the structure of the resulting parameter estimations. We assumed that the real data corresponds to the solution of model Eq. with parameters in Tables 2 and 3 which are randomly perturbed according to Eq. (8) with errors *ϵ*_*i*_ and *ϵ*_*j*_ assumed to be uniformly distributed with mean 0 and standard deviation 0.5. We produced two data sets. The first data set, which assumes data has been is collected daily from day 0 to day 12 post infection is

**Experiment 1:** 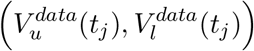 for *t*_*j*_ = {1, …, 12}.

The second data set, which assumes data is collected from day 7 to day 27 post infection is

**Experiment 2:** 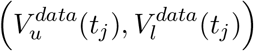 for *t*_*j*_ = {7, …, 27}.

Since the practical identifiability is a local property of the parameters, we used the priors for

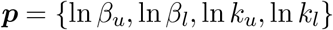

given in Table 3 and the priors for the rest of the parameters as in Table 2, to generate prediction graphs of the within-host model Eq. (7). The model”s predictive posterior distribution for all patients’ URT-LRT viral data for Experiments 1 and 2 are presented in Figure 6 together with grey areas for the 50% and 95% posterior regions (see also Figure S4). As expected, we observe wider model prediction ranges (gray regions) in the second phase decay for experiment 1 and in the expansion and peak areas for experiment 2, where data is scarce.

**Figure 6:**
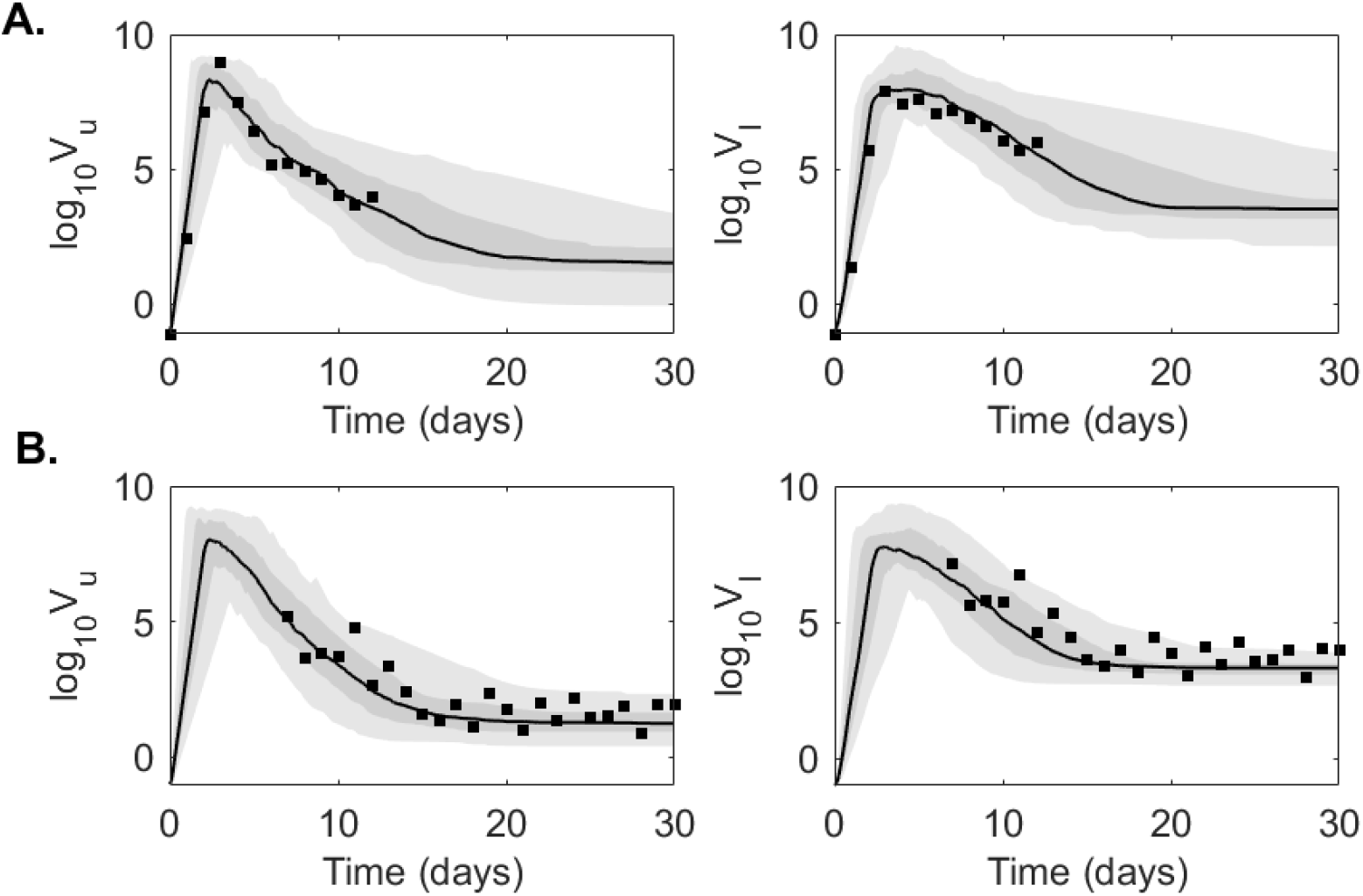
Virus dynamics obtained from fitting within-host model Eq. (7) to URT virus titer and LRT virus titer in (A.) Experiment 1 and (B.) Experiment 2. The grey bars represent 50% and 95% posterior regions.

To determine whether practical identifiability is lost in each experiment we created parameter histograms for each parameters (see Figure 7 and supplementary Figure S3). When data samples at the expansion stages of the infection are collected (as in Experiment 1), the LRT infectivity parameter *β*_*l*_ follows a normal distribution (see Figure 7A, left panel, blue bars). This results are validated by the corresponding dual parameter scatter plots (see Figure 8A). In contrast, when the data at the expansion stages of the infection is sparse (as in Experiment 2), the LRT infectivity parameter *β*_*l*_ follows a bimodal distribution (see Figure 7A, right panel, blue bars). This results are observed in the corresponding scatter plots, where we see bimodal clustering involving not just parameter *β*_*l*_, but involving parameter *β*_*u*_ as well (see Figure 8B). These results can be slightly improved when we consider that *β*_*u*_ and *β*_*l*_ follow lognormal distributions (see Figure 7B, right panel, blue bars). This suggests that the practical unidentifiability that appeared in the population data might be fixed by collecting data at the early stages of infection.

**Figure 7:**
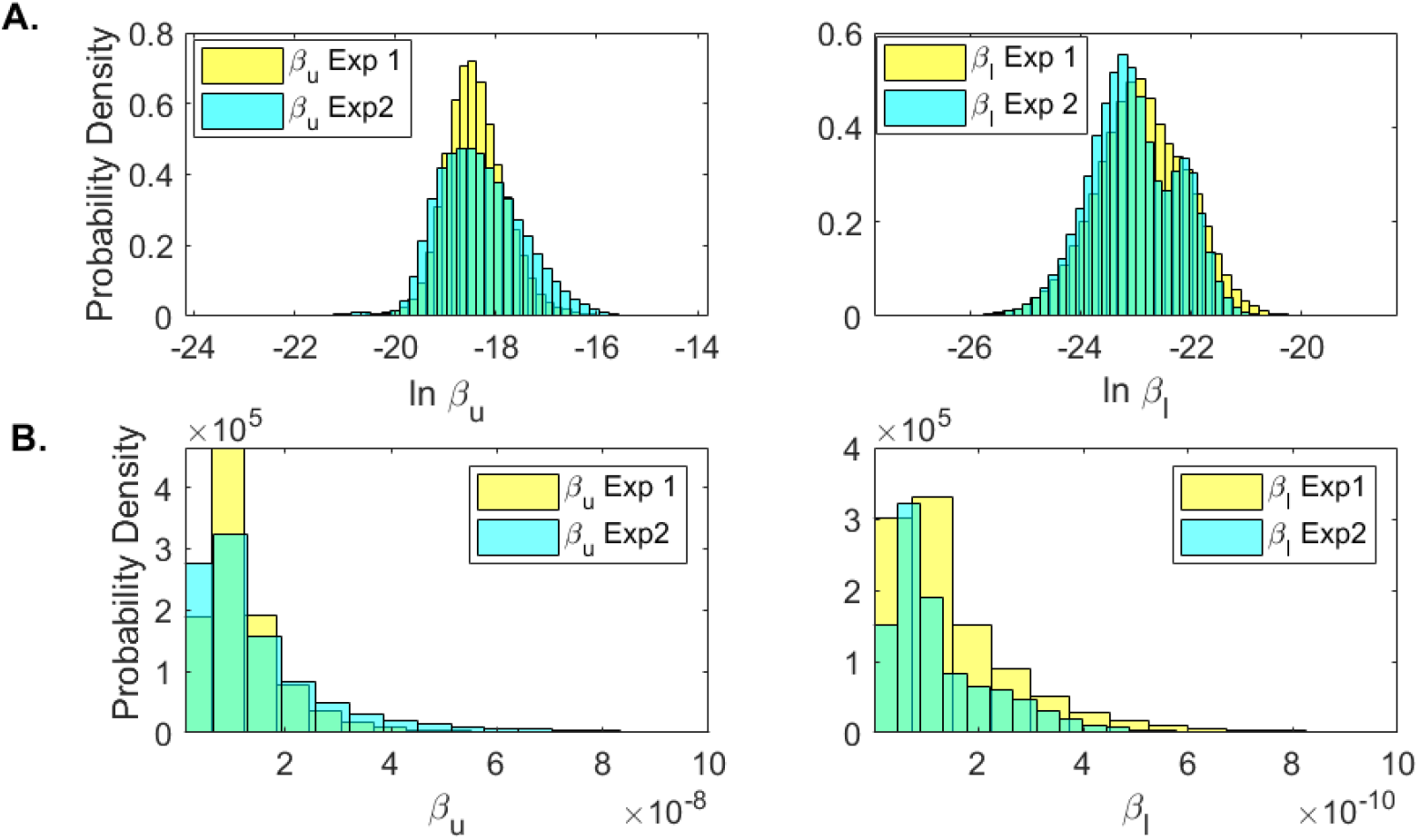
Histograms for *β*_*u*_ and *β*_*l*_ for Experiment 1 (yellow) and Experiment 2 (blue) when (A.) ln *β*_*u*_ and ln *β*_*l*_ are assumed to be normally distributed; and (B) *β*_*u*_ and *β*_*l*_ are assumed to be lognormally distributed.

**Figure 8:**
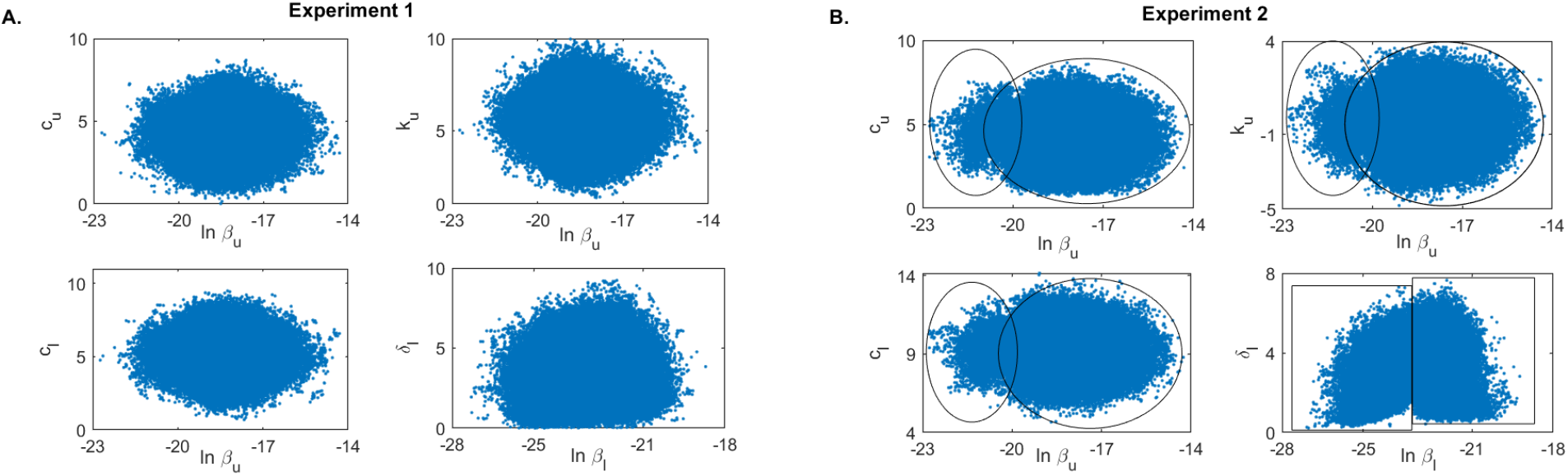
Scatter plots for (A.) Experiment 1 and (B.) Experiment 2. Parameters ln *β*_*u*_ and ln *β*_*l*_ are assumed to be normally distributed.

## Discussion

In this study, we developed a within-host mathematical model of SARS-CoV-2 infection that connected the virus kinetics in the upper and lower respiratory tracts of infected individuals and used it to determine the tract specific viral parameters. We removed viral production rates, to ensure structural identifiability, and fitted the rescaled model Eq. (7) to published longitudinal throat swabs and sputum titers in a single individual and in the entire population from SARS-CoV-2 infection study [44]. We estimated nine unknown parameters using an MCMC Bayesian fitting approach [19]. To avoid over fitting, we determined best estimates in a single patient (for which we have 26 data points) and in the entire population (for which we have 201 data points). We found shorter virus life-spans in LRT compared to viral URT, 2-3 hours compared to 5.7-8.5 hours. Our LRT estimates are similar to the fixed (and non-tract specific) virus life-span of 2.4 hours used by Ke *et al*. [24] and the estimated (and tract specific) life-span of 1.2 hours in Wang *et al*. [42], but longer than the 10 hours seen in influenza and used by Hernandez *et al*. [22]. The between tracts differences may suggest the presence of additional immune mediated viral clearance in the LRT. We found similar infected cells life-span between the two tracts, with a range of 4.2-8 hours, shorter than in other studies [22, 24]. Lastly, the mean URT basic reproductive number for the entire population, 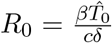, equals 17.4, higher than in [24]. While we assumed two-way viral shedding between tracts, data fitting suggested higher virus shedding from upper and lower respiratory tracts than the other way around, consistent with other studies [24].

Interestingly, we found that the estimated LRT infectivity rate parameter follows a bimodal distribution when the model was fitted to the entire population data. We attributed this behavior to practical non-identifiability. Practical identifiability is observed when the measured data is contaminated with noise. We have inherently accounted for noisy data by combining viral measurements from nine patients with different viral profiles. We investigated several ways for improving practical identifiability of this parameter and found that both estimating the logaritmic value of this parameter ln *β*_*l*_ and assuming log-normal distributions for some parameters improves the accuracy of our estimates.

Most importantly, it has been reported that practical identifiability can be achieved by adding pertinent data measurements that can help improve the identity of unknown parameters [14, 43]. Such a process, known as optimal experimental design, aims to obtain additional information about a system through the addition of new measurements. Since in system Eq. (7) the non-practically identifiable infectivity parameter *β*_*l*_ is responsible for the LRT dynamics early in the infection, we investigated whether the addition of early data contains the maximal information needed for improving its estimate. We created two virtual data sets, one in which data is collected daily for the first 12 days and one in which data is collected daily for 20 days, starting at day 7. We found that the infectivity rate *β*_*l*_ is bimodal and, hence, non-practically identifiable when data is missing during the first seven days of infection. The absence of early data leads to an underestimation of overall LRT viral titer in the first 14 days following infection (see Fig. S4). This may affect one’s ability for determining the best window for antiviral and immune modulation interventions [9]. Moreover, it will provide a underestimate for the period of maximum infectiousness [21], which may affect recommendations for quarantine and isolation [13]. Hence, the existence of data measurements before and/or at symptoms onset is crucial for best parameter estimation and model prediction when considering noisy population data.

Our study has several limitations. First, we considered a density dependent clearance term for the URT virus that saturates at around 1-2×10^3^ RNA copies per ml, in order to explain the viral RNA persistence in the LRT at 30 days following infection reported in the Wolfle *et al*. [44]. While in public health setting a SARS-CoV-2 diagnostic is determined by PCR assays, long-term RNA levels are not a reliable measurement of infectiousness, with the measured RNA values indicating the presence of genomic fragments, immune-complexed or neutralised virus, rather than replication-competent virus [1,21,34]. Further work is needed to separate the presence of infectious versus noninfectious viral RNA in the lower respiratory tract. Secondly, we did not consider an eclipse phase in the virus infectiousness (usually assumed to be around 6 hours [21, 24]). This simplification may be the leading reason for larger estimates for the death rate of infected cell in our study compared to other studies [22, 24]. Thirdly, due to the novelty of the model, we have no information on parameter priors. Therefore, we fitted the within-host model to the patient A and population data, and used those estimates as a mean in the prior distributions. However, since the resulting means fall within ranges observed for other acute infections [2–4, 6, 31, 32], and since we consider large standard deviations around the prior means, we are confident that we are covering a large search space that does not exclude viable outcomes.

In conclusion, we have developed a within-host model of SARS-CoV-2 infection in the upper and lower respiratory tracts, used it to determine pertinent viral parameters, and suggested the optimal experimental designs that can help improve the model predictions. These techniques may inform interventions.

## Data Availability

Code is available upon request.

## Acknowledgements

SMC acknowledges support from National Science Foundation grants No. DMS-1813011 and DMS-2051820 and by a Virginia Tech Center for Emerging, Zoonotic, and Arthropod-borne Pathogens (CeZAP) seed grant. NT acknowledges partial support from National Science Foundation grant DMS-1951626.

---

**Figure S1:**
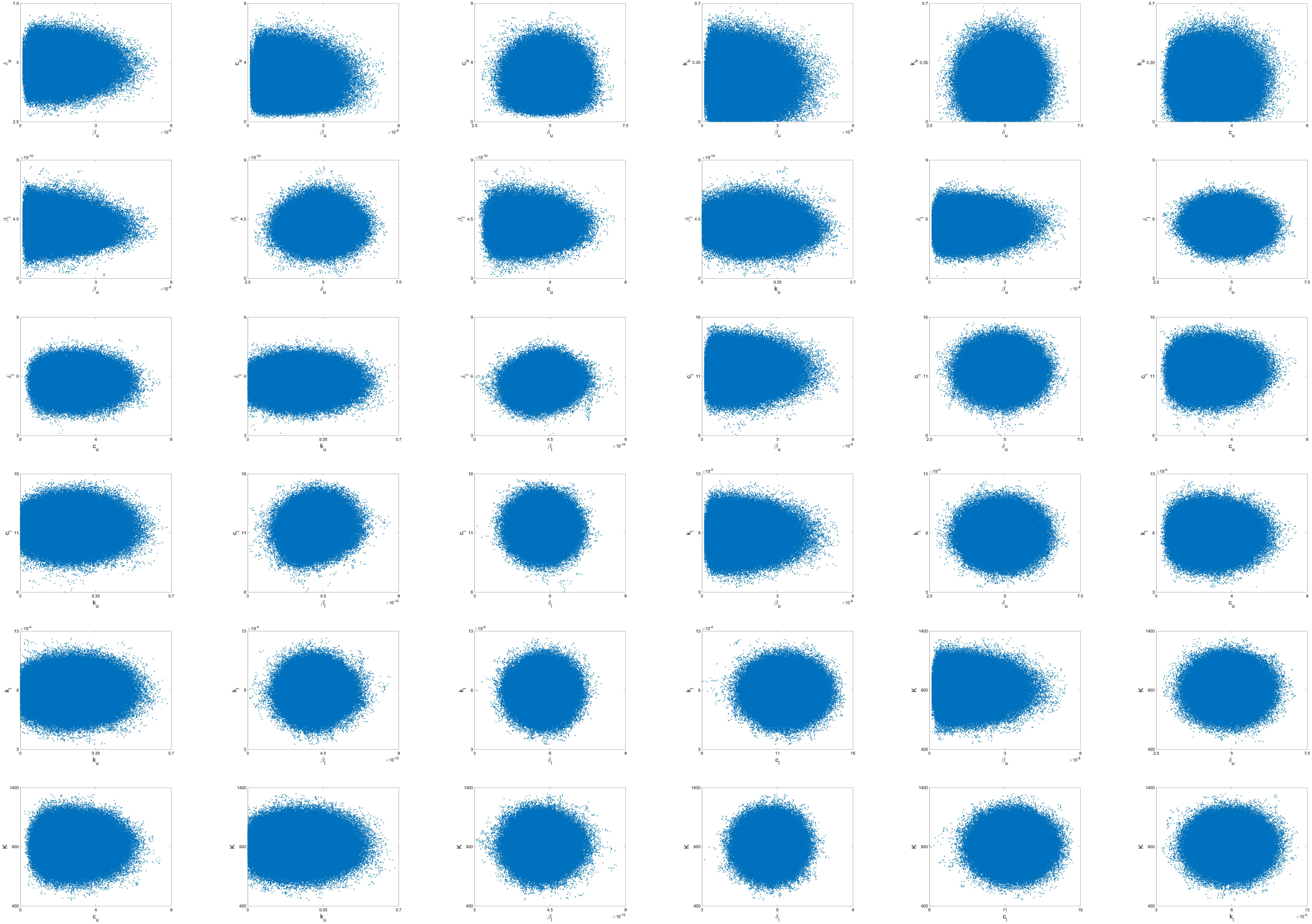
Two-parameter scatter plots for patient A. We sampled the parameter space *N* = 10^6^ times.

**Figure S2:**
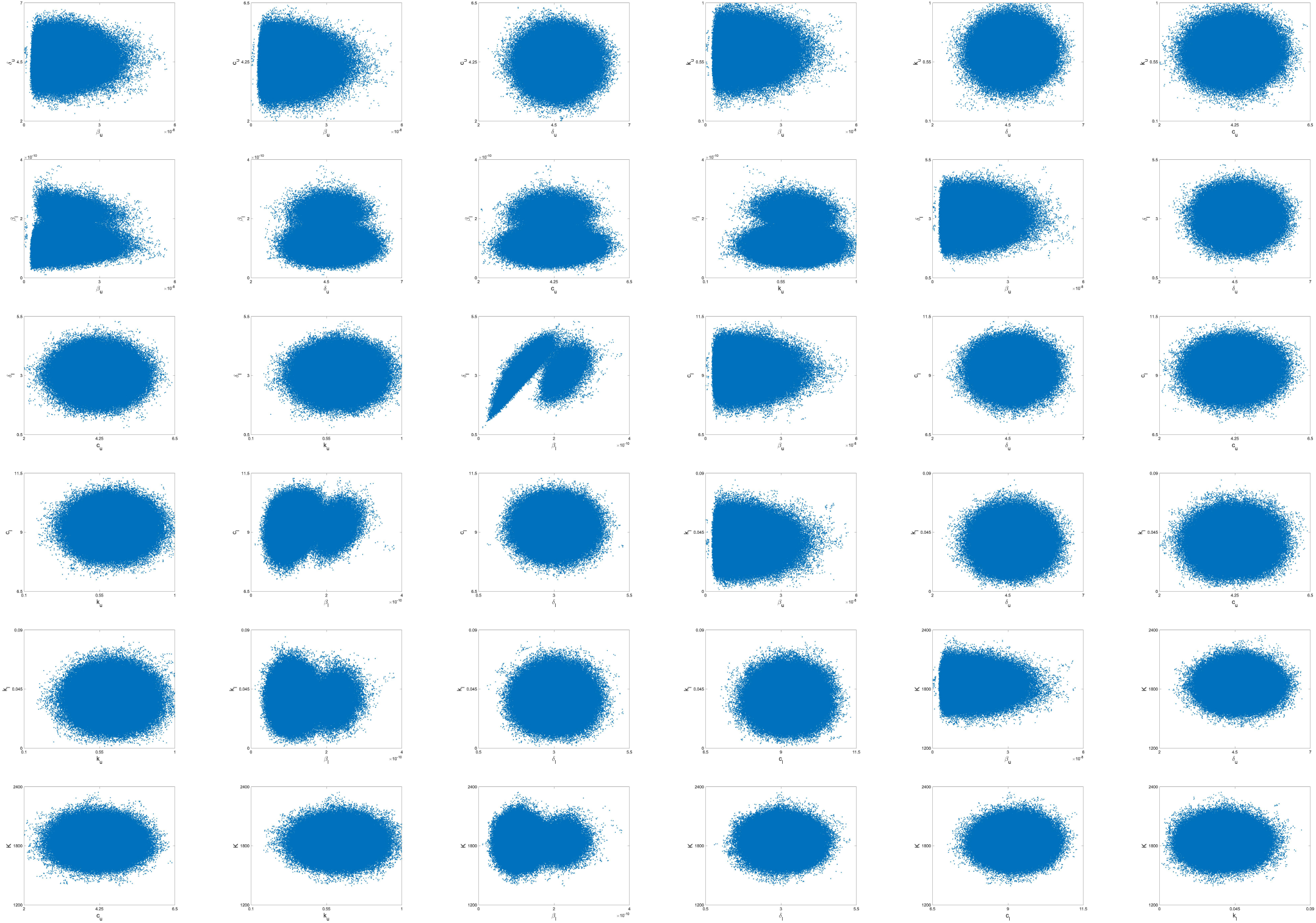
Two-parameter scatter plots for the total population. We sampled the parameter space *N* = 10^6^ times.

**Figure S3:**
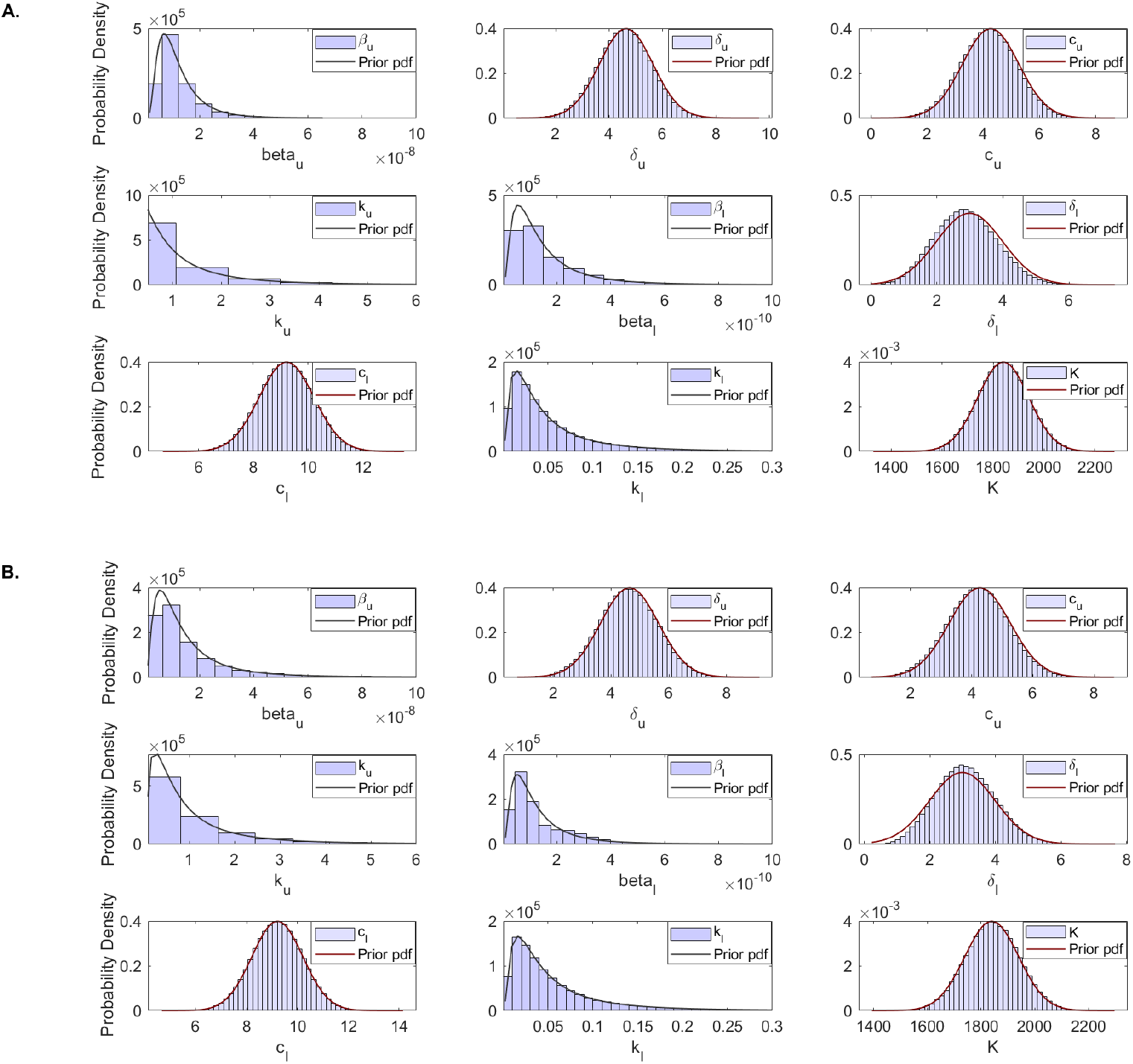
Histogram of estimated parameter distributions from fitting model Eq. (7) to virtual URT virus titer and LRT virus titer data in: (A.) Experiment 1 and (B.) Experiment 2. Parameters *β*_*u*_, *β*_*l*_, *k*_*u*_, *k*_*l*_ were considered lognormal distributed. All other parameters were considered normally distributed.

**Figure S4:**
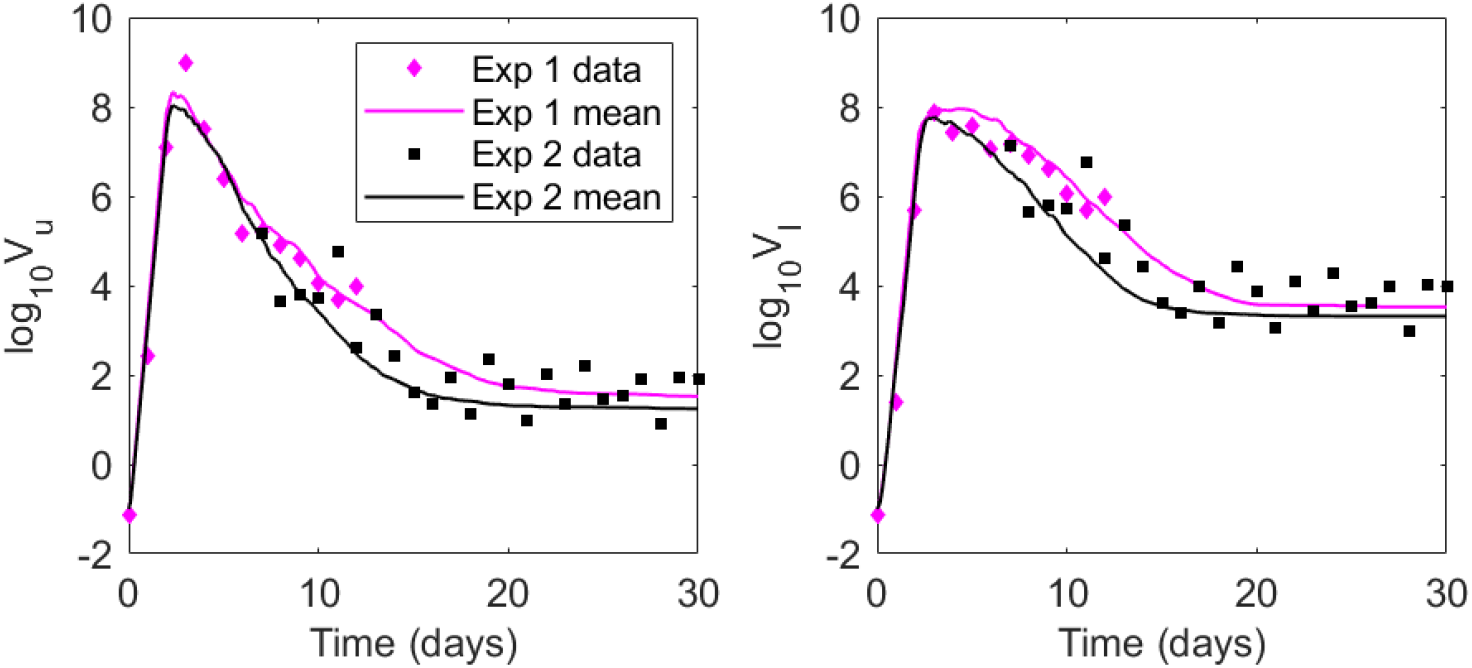
Virus dynamics obtained from fitting within-host model Eq. (7) to (left panel) URT virus titer and (right panel) LRT virus titer in Experiment 1 (magenta) and Experiment 2 (black).

